# A completeness indicator of gestational and congenital syphilis information in Brazil

**DOI:** 10.1101/2022.05.29.22275727

**Authors:** Guilherme Lopes de Oliveira, Andrêa JF Ferreira, José Guilherme Santana, Raquel Martins Lana, Andrey Moreira Cardoso, Carlos Teles, Rosemeire L. Fiaccone, Rosana Aquino, Maria Auxiliadora Santos Soares, Enny S. Paixão, Idália Oliveira dos Santos, Leonardo Salvi, Mauricio L. Barreto, Maria Yury Ichihara

## Abstract

**Objective:** To evaluate the quality of information on gestational syphilis (GS) and congenital syphilis (CS) on the Notifiable Diseases Information System (SINAN-Syphilis Brazil) by compiling and validating completeness indicators between 2007 and 2018.

**Materials and methods:** Overall, care and socioeconomic completeness scores were compiled based on selected variables, using weights assigned by specialists. The completeness scores were analysed, considering the region and area of residence, pregnant race/colour, and year of case notification. Pearson’s correlation coefficients were used to validate the scores obtained by the weighted average method, compared with the values obtained through principal component analysis (PCA).

**Result:** Most of the variables selected presented a good or excellent degree of completeness for GS and CS, except for clinical classification, pregnant woman’s level of education, partner’s treatment, and child’s race/colour, which were classified as poor or very poor. The overall (89.93% versus 89.69%) and socioeconomic (88.71% versus 88.24%) completeness scores for GS and CS, respectively, were classified as regular, while the care score (GS-90, 88%, and CS-90, 72%) was good, although there were improvements over time. Differences in the overall, care and socioeconomic completeness scores according to region, area of residence, and ethnic-racial groups were reported for syphilis notifications. A strong linear correlation (>0.90) was observed between the completeness scores estimated through the weighted average method and PCA.

**Conclusion:** Improvements in the completeness of GS and CS notifications have been observed in recent years, highlighting the variables which form the care score, compared to the socioeconomic scores, despite differences between regions, area of residence, and ethnic-racial groups. The weighted average was a viable methodological alternative easily operationalised to estimate data completeness scores, allowing routine monitoring of the completeness of gestational and congenital syphilis records.

## Introduction

Problems with the quality of data registered on Brazilian Health Information Systems (HIS) remain, although undeniable progress has been made in recent years^1^. Different parameters have been used to evaluate the quality of health information, including completeness^2,3^, defined as the degree of variable completion on HIS, a thermometer of the quality of information generated by these systems^3^.

The incompleteness of information from HIS impairs the accuracy of records, affecting the reliability of the information, the adequate knowledge of the health-disease process, and the epidemiological patterns of events^4,5^. The capacity to plan appropriate actions to prevent and control disease and disorders within the scope of epidemiological surveillance, integrated into those of care, is reduced^6^. Then, HIS completeness is essential to promote and protect the population’s health and the appropriate planning and management of resources, including investments for the continuous improvement of these systems by the Ministry of Health^2,7^.

Syphilis is among the diseases of great relevance in public health, and, therefore, we need qualified information for its monitoring. As a secular disease and historically neglected, syphilis is caused by the bacterium *Treponema pallidum* and transmitted mainly sexually and vertically^8,9^. World Health Organization (WHO) estimates that approximately 12 million new syphilis cases occur annually, mainly in low-and-middle-income countries^9^. During pregnancy, labour, and postpartum phases, the mother-baby binomial has been the focus to track maternal syphilis during prenatal care, preventing vertical transmission of syphilis.

In Brazil, approximately 61,441 cases of gestational syphilis (GS) and 22,065 cases of CS, with a GS detection rate of 20.8 and CS incidence rate of 8.9 per 1,000 live births^10^, were reported in 2021. CS was only included on the national list of diseases requiring mandatory notification in 1986, GS and acquired syphilis were added in 2005 and 2010, respectively^6,11^, being reported through individual notification records (INR) on the Notifiable Diseases Information System (SINAN). Filling INR fields, including those not mandatory, is recommended following confirmation of a syphilis case^6^. Analysing the information generated by GS and CS notification records is fundamental to monitoring diseases indicators and ensuring the improvement of prevention, control, and care, besides indicating strategies to enhance its completion quality.

Few national studies have evaluated the completeness of SINAN-Syphilis information^7,12,13^ and the factors that influence its variation, such as region, year of notification, area of residence, and race/colour of the individual, as has been evidenced in other studies with HIS in Brazil^2,7,14^. There is also a lack of valuable indicators which are easily operationalised, which enable health managers and professionals to monitor the completeness of the information registered on HIS. This study aims to evaluate the quality of the information by compiling and validating SINAN-Syphilis completeness indicators in Brazil between 2007 and 2018.

## Materials and Methods

This descriptive and analytical study evaluated the degree of completeness of the variables in the GS and CS records available on the Ministry of Health SINAN-Syphilis between 2007 and 2018^6^. We build binary indicators and completeness scores for the overall variables selected and for those classified as socioeconomic or related to care. Completeness degree was classified according to Romero and Cunha^15^ as follows: excellent (> 95%), good (90%- 95%), regular (80%-90%), poor (50%-80%) and very poor (≤ 50%).

The following variables were selected: **GS** – pregnant woman’s age range; area of residence; level of education, race/colour; gestational age; qualitative results of treponemal and non-treponemal tests (VDRL); treatment regime; clinical classification and partner’s treatment; and **CS** - age range; child’s sex and race/colour; area of residence; mother’s level of education and race/colour; qualitative results of VDRL for the child; case evolution; attending prenatal care; treatment regime; time of diagnosis and partner’s treatment.

### 1. Building the completeness indicators and scores

#### (a) Binary completeness indicator

to evaluate the individual completion of each variable, the proportion of individual notification records with complete information was obtained, considering the indicator of the presence (1) or absence of information, information ignored, or inconsistent (0) in the selected variables.

#### (b) Completeness scores

to evaluate the completion of a set of variables, the weighted average method was used, based on the values of the binary indicators defined in (a) and weights defined on a scale from 1 to 4, varying from the lowest to the highest importance, respectively. The weights were assigned independently by specialists (epidemiologists and health professionals), considering their conception of the relevance of each variable to analyse epidemiological patterns from the perspective of the prevention and control of GS and CS. Subsequently, these weights were evaluated and agreed upon by the study researchers for their use in the estimates of completeness scores (Table 1). The overall score considers all variables selected, while the socioeconomic and care scores consider the variables classified as socioeconomic and care, respectively. The three scores vary between 0 and 1, and the higher the score value, the better the completeness of the set of variables considered. The completeness scores were analysed according to the year of case notification, region, area of residence, and pregnant woman’s race/colour.

**Table 1.**
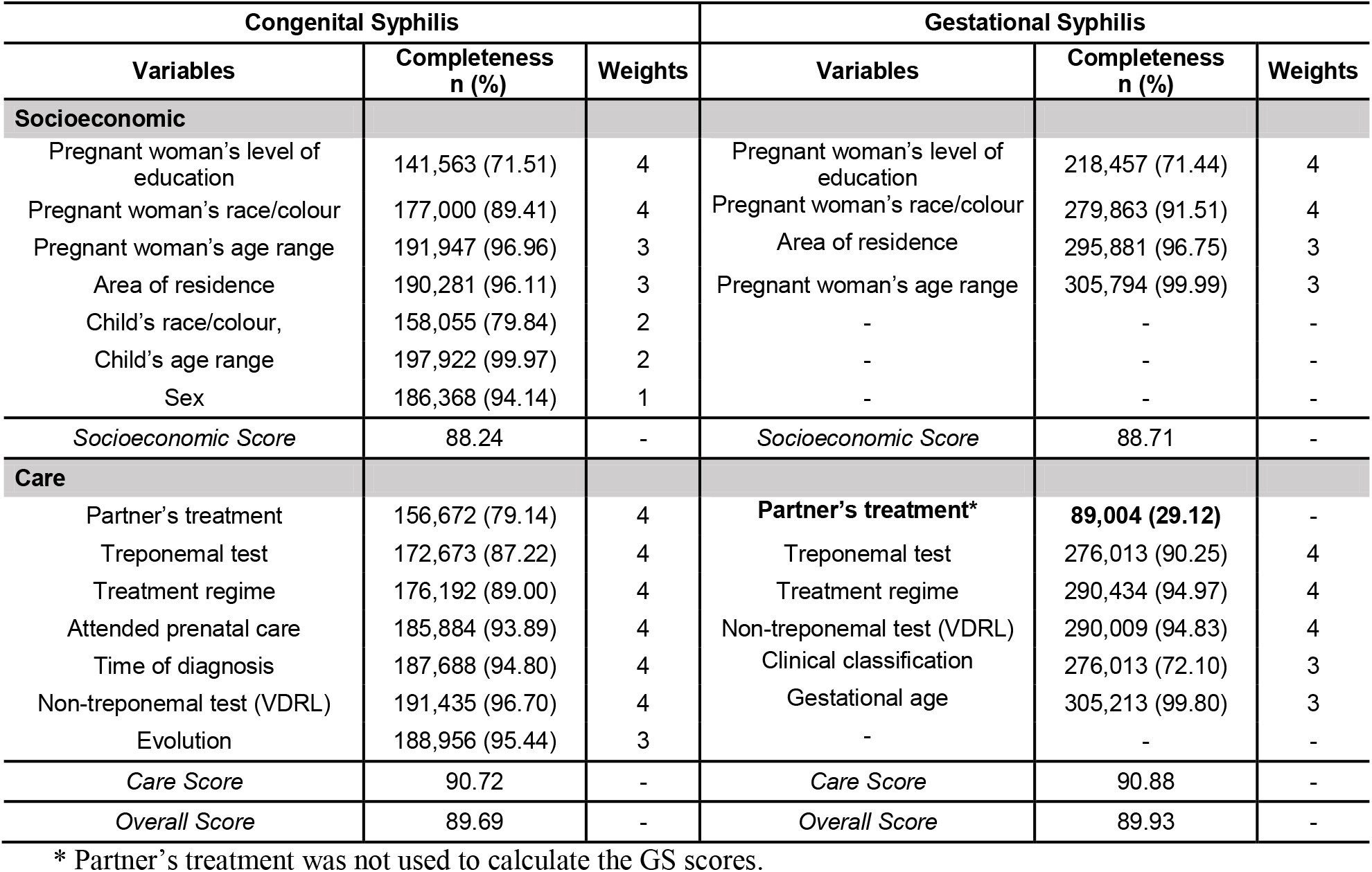
Analysis of the completeness of the overall, socioeconomic and care variables and scores. SINAN-Syphilis, 2007-2018.

| Congenital Syphilis |  |  | Gestational Syphilis |  |  |
| --- | --- | --- | --- | --- | --- |
| Variables | Completeness<br>n (%) | Weights | Variables | Completeness<br>n (%) | Weights |
| **Socioeconomic** |  |  |  |  |  |
| Pregnant woman's level of education | 141,563 (71.51) | 4 | Pregnant woman's level of education | 218,457 (71.44) | 4 |
| Pregnant woman's race/colour | 177,000 (89.41) | 4 | Pregnant woman's race/colour | 279,863 (91.51) | 4 |
| Pregnant woman's age range | 191,947 (96.96) | 3 | Area of residence | 295,881 (96.75) | 3 |
| Area of residence | 190,281 (96.11) | 3 | Pregnant woman's age range | 305,794 (99.99) | 3 |
| Child's race/colour, | 158,055 (79.84) | 2 | - | - | - |
| Child's age range | 197,922 (99.97) | 2 | - | - | - |
| Sex | 186,368 (94.14) | 1 | - | - | - |
| *Socioeconomic Score* | 88.24 | - | *Socioeconomic Score* | 88.71 | - |
| **Care** |  |  |  |  |  |
| Partner's treatment | 156,672 (79.14) | 4 | **Partner's treatment\*** | **89,004 (29.12)** | - |
| Treponemal test | 172,673 (87.22) | 4 | Treponemal test | 276,013 (90.25) | 4 |
| Treatment regime | 176,192 (89.00) | 4 | Treatment regime | 290,434 (94.97) | 4 |
| Attended prenatal care | 185,884 (93.89) | 4 | Non-treponemal test (VDRL) | 290,009 (94.83) | 4 |
| Time of diagnosis | 187,688 (94.80) | 4 | Clinical classification | 276,013 (72.10) | 3 |
| Non-treponemal test (VDRL) | 191,435 (96.70) | 4 | Gestational age | 305,213 (99.80) | 3 |
| Evolution | 188,956 (95.44) | 3 | - | - | - |
| *Care Score* | 90.72 | - | *Care Score* | 90.88 | - |
| *Overall Score* | 89.69 | - | *Overall Score* | 89.93 | - |

\* Partner's treatment was not used to calculate the GS scores.

### 2. Validating the completeness scores

The validation was based on comparing the scores obtained by the weighted average method with those obtained through principal component analysis (PCA), a multivariate statistical technique used to reduce dimensionality, index generation (scores), and clustering of individuals or variables. The tetrachoric correlation matrix of the binary completeness indicators was used to extract the principal components^16^. The binary indicators of the variables with excellent completeness (> 95%) were not considered in the analysis due to the low variability between the categories, and the “partner’s treatment” variable was disregarded in the CS analysis due to the high percentage of data either missing or ignored. We calculated the PCA scores for each record using the regression method^17^. The average of scores associated with the first two principal components extracted in the analysis was considered for the overall score. For socioeconomic and care scores, only the first principal component was considered. Lastly, we analyse the relationship between scores calculated by the weighted average and PCA methods through scatter plots and estimate Pearson’s linear correlation coefficient. R software, version 3.6.0, and the psych package were used in the PCA^18^.

#### Ethical Aspects

This study used anonymised administrative health data provided by the Ministry of Health, exempting submission to the Ethics and Research Committee.

## Results

305,891 GS and 197,974 CS cases were notified between 2007 and 2018, registering an increase of 9.1 and 5.3 times, respectively, in notification cases in 2018. Notification of GS was higher in the Southeast (44.8%) and Northeast (21.3%) regions, followed by the South (14.4%), North (10.8%), and Central-West (8.6%). A similar pattern was found for CS. Considering the area of residence of GS notifications, 88.3% occur in the urban area, 7.7% in rural, 0.7% from the peri-urban area and 3.2% of the notifications did not fill out this information. Most CS notifications (89.3%) were from urban areas, 6% from rural and 0.8% from peri-urban areas, and 3.9% of the cases did not include this information. The prevalence of GS cases was concentrated among women who self-declared mixed/brown (48.3%), white (29.3%), black (12.3%), asian descendant (0.9%) and indigenous (0.8%), with information on race/colour being absent in 8.5% of the notifications. For CS, 48.9% of case notifications was mixed/brown, 25.4% white, 5.0% black, 0.4% indigenous, 0.24% asian descendant, and 20.1% of notifications were without any information on race/colour.

Analysis of the completeness of GS records, measured by the binary indicator (Table 1), was classified as excellent (> 95%) for the pregnant woman’s age range, area of residence, and gestational age; good (90%-95%) for the pregnant woman’s race/colour, qualitative results of treponemal test, VDRL test and treatment regime; poor (50%-80%) for the pregnant woman’s level of education and clinical classification; and very poor (<50%) for the partner’s treatment information. The CS notification records presented excellent completeness (>95%) for the mother and child’s age range variable, area of residence, qualitative results of VDRL test, and clinical evolution; good (90%-95%) for child’s sex, prenatal care, and the time of diagnosis; regular (80%-90%) for the mother’s race/colour, treponemal test and treatment regime; and poor (50%-80%) for the mother’s level of education, child’s race/colour, and partner’s treatment.

Moreover, overall and socioeconomic completeness scores for GS (89.93% and 88.7%) and CS (89.69% and 88.71%) were classified as regular (Table 1), while the care score was good (GS-90.88% and CS-90.72%) over the years.

Between 2007 and 2015 for Brazil (Figure 1), the overall completeness score was evaluated as regular for GS (87.7% in 2007 to 89.9% in 2009) and CS (85.5% in 2007 to 89.9% in 2014) and good from 2016. For both diseases, the socioeconomic score was also classified as regular over the years, with a growing tendency to improve completeness between 2011 and 2017 for GS (87.5% and 89.8%), and between 2007 and 2017 for CS (85.5% and 89.4%). The care score was classified as regular for GS between 2007 and 2012 and good in the following years, while for CS, it remained regular until 2015, with a gradual increase in the completeness scores in the subsequent years. We highlighted that, from 2008, the care score was higher than the socioeconomic score for both diseases. The North and South Brazilian regions presented, over time, improvement in completeness scores for GS and CS, especially for socioeconomic scores (Figure 1). This improvement occurred in the Central-West and Southeast regions for GS, while for CS, it was more evident in the Southeast and Northeast regions.

**Figure 1.**
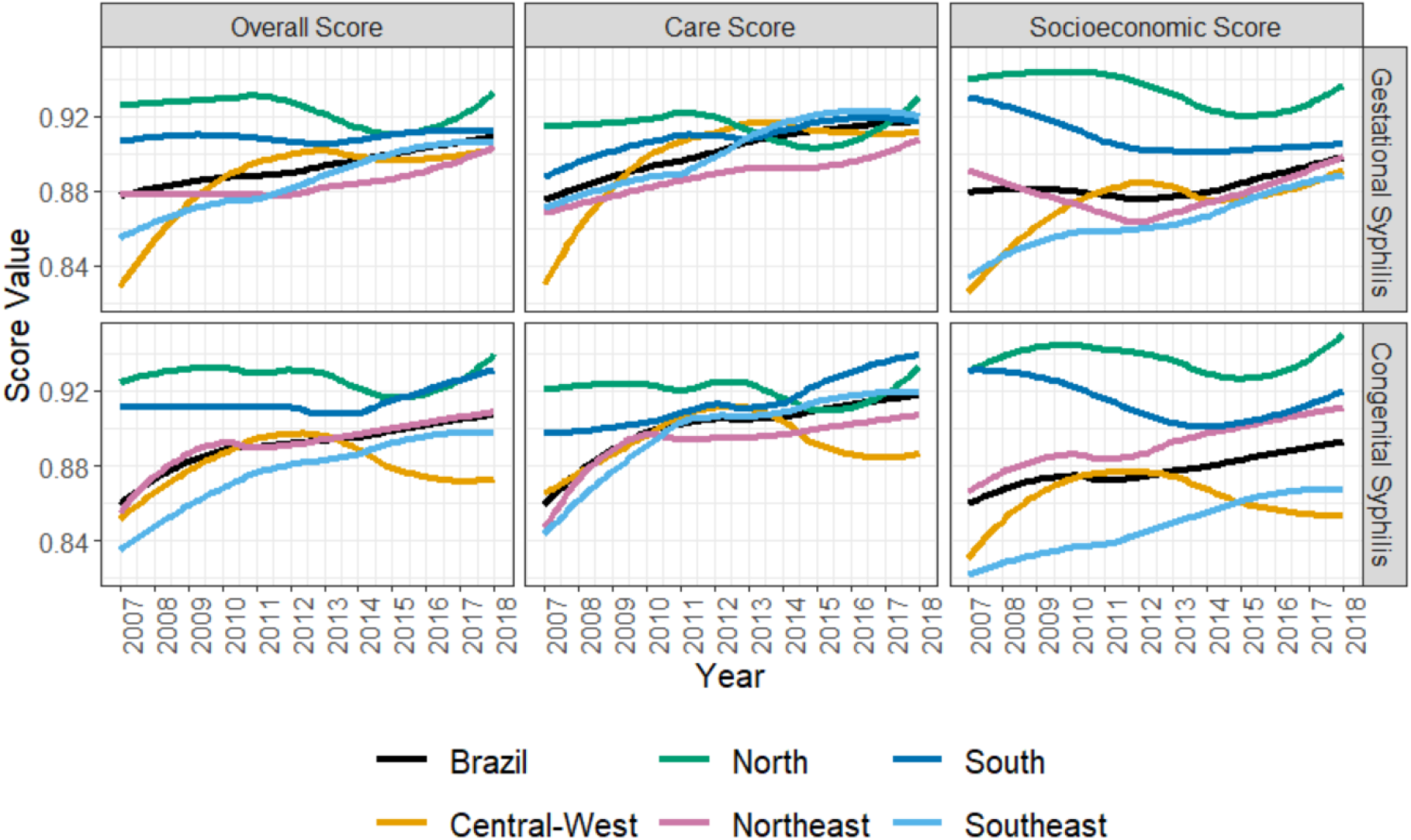
Analysis of completeness scores according to the year of case notifications and Brazilian regions SINAN-Syphilis, 2007-2018.

It is essential to highlight the regional differences observed in the gestational and congenital completeness scores (Figure 2): the overall and socioeconomic scores were classified as good in the North (GS-92.2% and CS-92.6% and GS-93.1% and CS-93.7%, respectively) and South regions (GS-91% and GS-90.5% and CS-92% and CS-91.2%, respectively), and regular in the other regions. The care score for GS was classified as good in all regions (North-91.4%; Central-West-90.5%; Southeast-91.3%; and South-91.5%), except for the Northeast region (89.4%), classified as regular. For CS, the care score was classified as good for the North (91.9%), Southeast (90.8%), and South regions (92.5%), and regular for the Northeast (89.7%) and Central-West (89.1%) regions.

**Figure 2.**
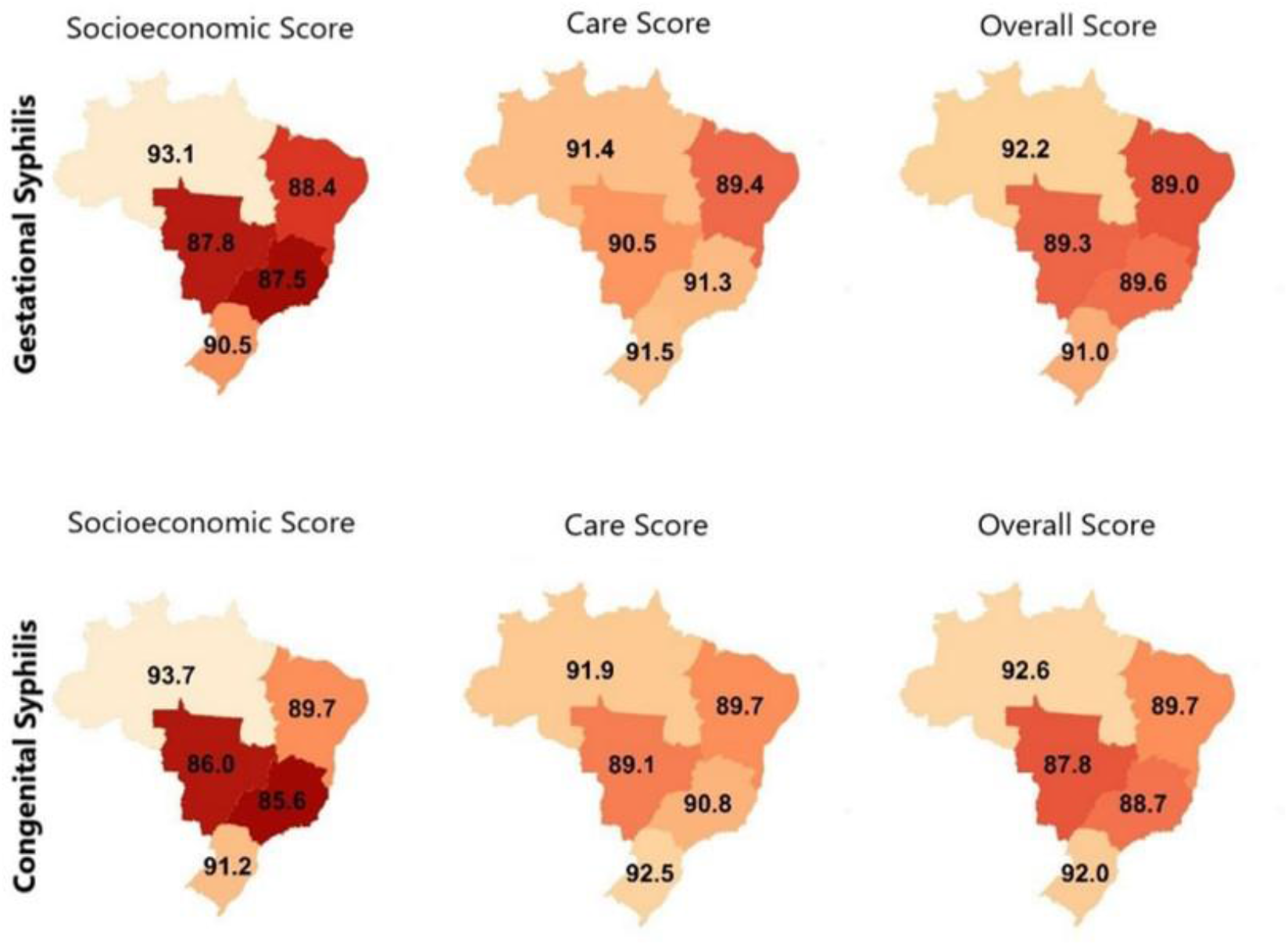
Analysis of completeness scores according to the region of residence for gestational and congenital syphilis, SINAN-Syphilis, 2007-2018. Legend: The numerical values correspond to the score value observed in each region. The darker scores correspond to the lowest completeness scores and the lightest scores to the highest ones.

Gestational and congenital syphilis cases presented information for the area of residence in 96.8% and 96.1% of notification records, respectively (Figure 3). It was observed that the overall and care scores classifications were good in the urban (GS-90.4% and 91.1%; CS-90.1% and 91%), rural (GS-91.3% and 90.8%; CS-91.5% and 90.8%) and peri-urban area of residence (GS-90.6% and 90.5%; CS-90.5% and 91.1%). However, the socioeconomic score presented a good quality for GS in rural (92.0%) and peri-urban areas of residence (90.8%) but regular in the urban area (89.5%). For CS, the socioeconomic score was good for the rural area (99.2%) and regular for the other areas of residence (urban-88.9% and peri-urban-89.7%). However, when information about the area of residence was ignored (GS-3.2% and CS-3.9%), the classification of completeness scores worsened (overall and care scores were poor, while the socioeconomic score was regular).

**Figure 3.**
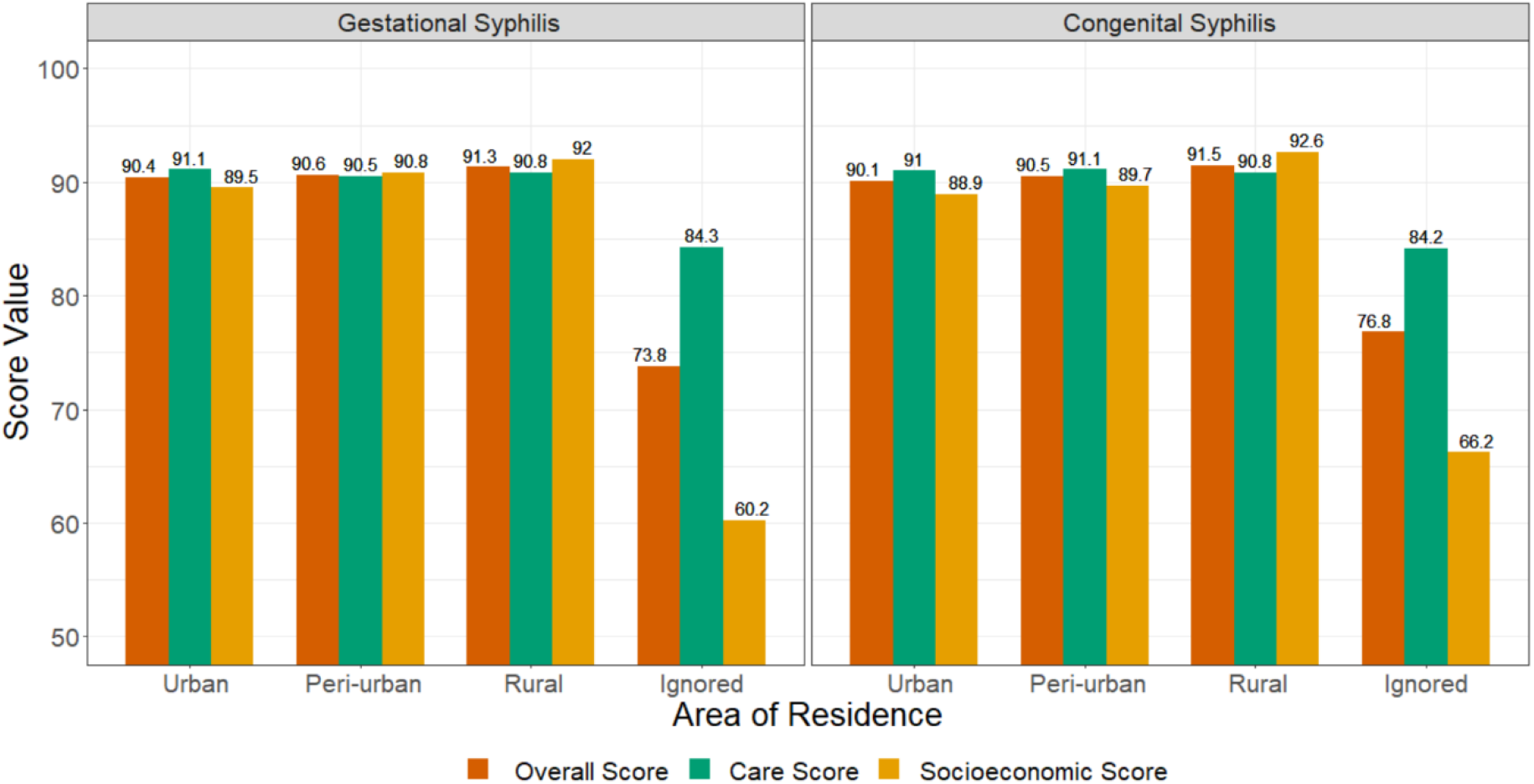
Analysis of completeness scores according to the area of residence of pregnant women for gestational and congenital syphilis, SINAN-Syphilis, 2007-2018.

Considering 91.5% and 79.85% of the GS and CS cases (Figure 4), respectively, for which information on race/colour was filled, we observed that the overall, socioeconomic and care completeness scores for GS were classified as good for all the racial categories (black, mixed/brown, indigenous, asian descendant, and white). For CS, the overall and socioeconomic scores presented good completeness for all the racial categories. In cases where race/colour information was ignored (GS-8.5% and CS-20.15%), all the completeness scores were classified as poor, except the socioeconomic score of GS records, which was regular.

**Figure 4.**
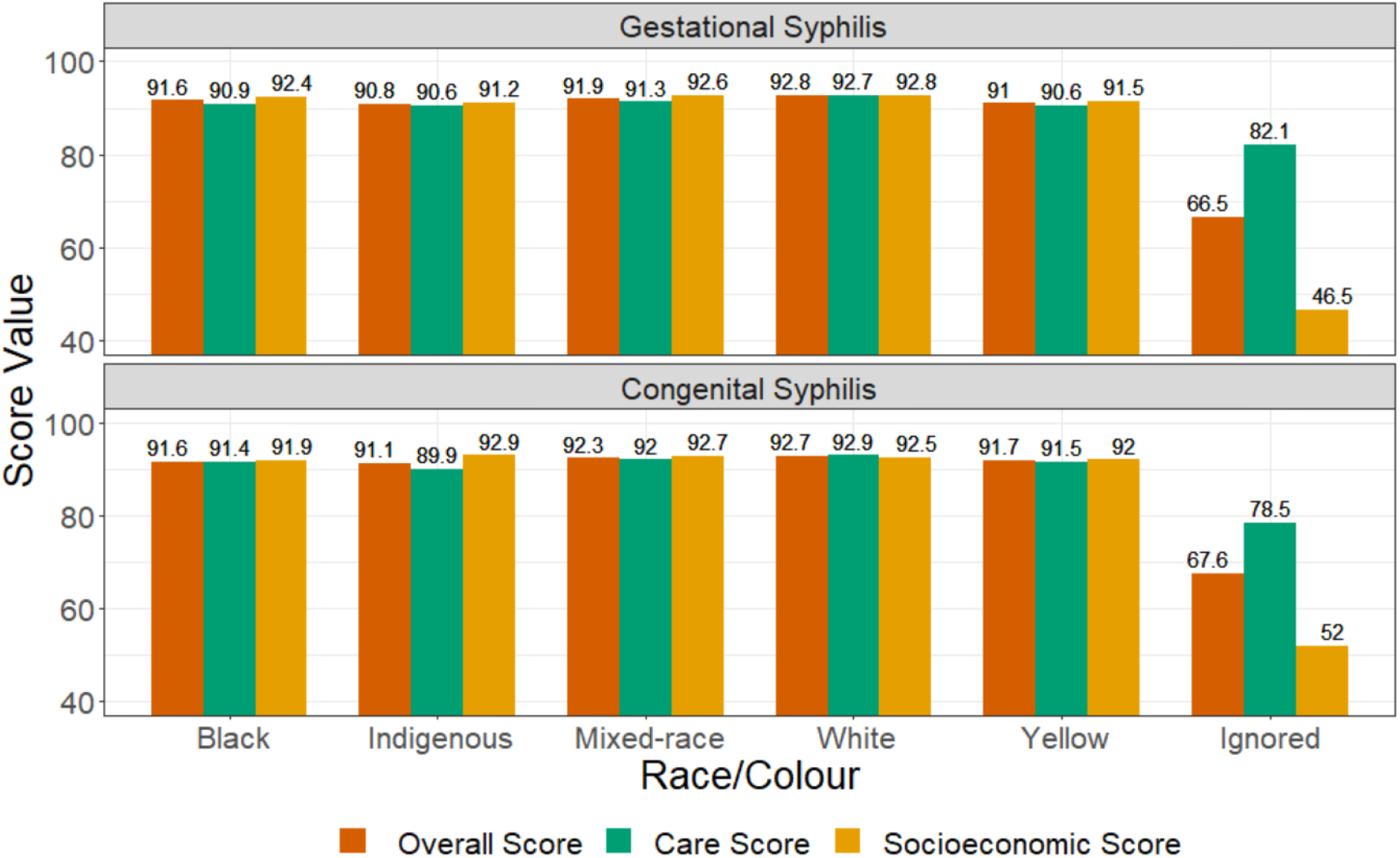
Analysis of completeness scores according to the pregnant woman’s race/colour for gestational and congenital syphilis, SINAN-Syphilis, 2007-2018.

Validation of the overall, socioeconomic and care scores were achieved by comparing them using the weighted average and PCA methods, and the result presented a strong linear correlation (>0.90) in the scatter plots (Figure 5). In the case of PCA, the explanation percentage for the principal components retained in the analysis varied between 47.15% and 63.30% for the six different scores calculated. The result obtained by both methods was similar.

**Figure 5.**
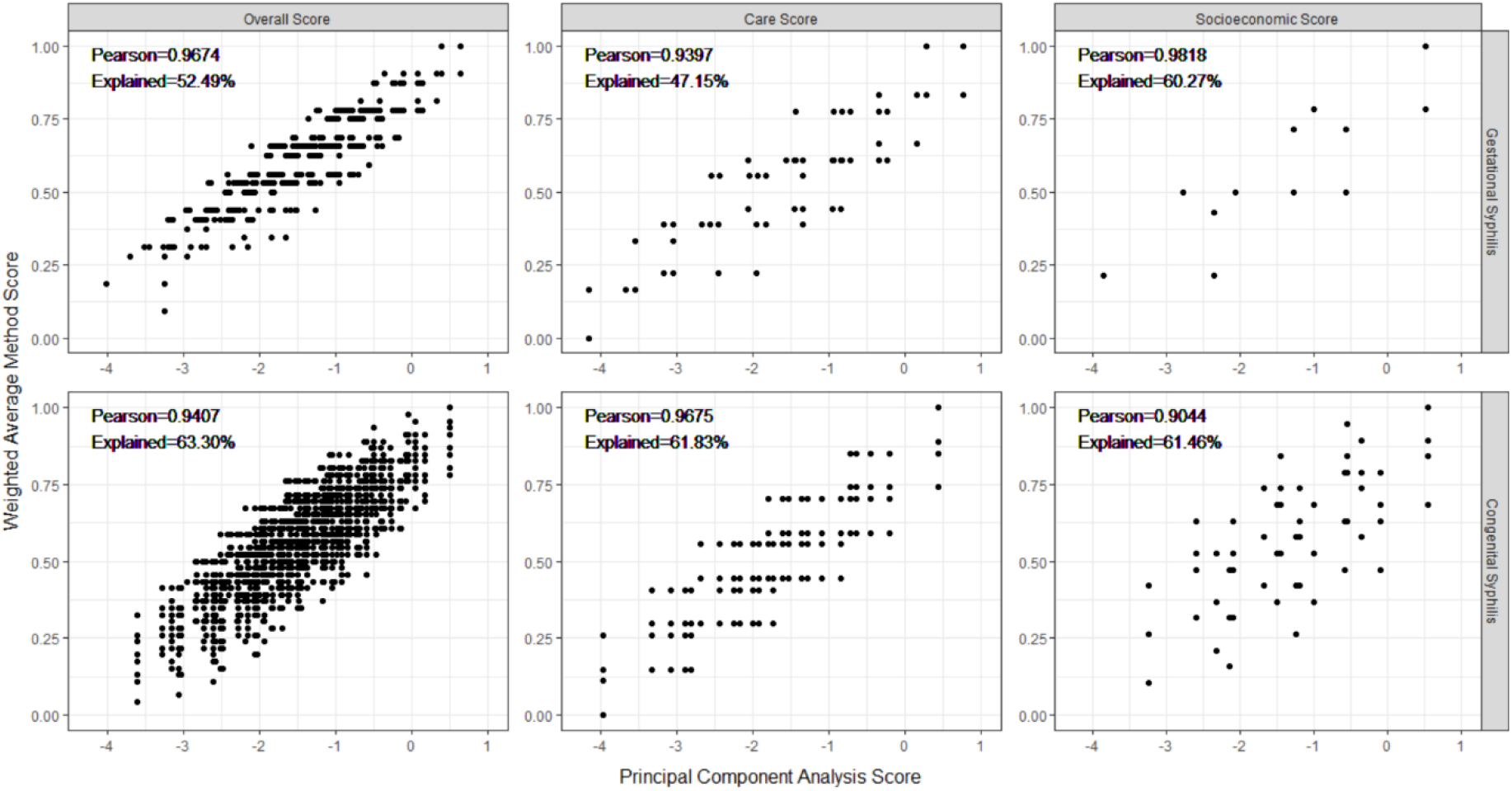
Comparison between the completeness scores obtained through the weighted average method and those obtained through principal component analysis, SINAN-Syphilis, 2007-2018. Legend: We have the GS comparisons on the upper line and the CS results on the lower line. The comparative graphs for the socioeconomic scores are presented in the left-hand column; the comparison of the care scores in the central column and the overall scores are compared in the right-hand column. The graphs present Pearson’s linear correlation coefficient (Pearson’s R). The explanation percentage obtained in the PCA is presented on the horizontal axis of the graphs.

## Discussion

The results indicate a good or excellent degree of completeness for most of the SINAN-Syphilis variables included in gestational and congenital syphilis notification records, except for clinical classification, pregnant/mother’s level of education, partner’s treatment, and child’s race/colour, which were classified as poor or very poor. The overall and socioeconomic completeness scores were similar for GS and CS records, both classified as regular, while the care score was classified as good and improved during the period studied. However, differences in scores completeness were verified according to region, area of residence, and ethnic-racial groups. These differences were also observed in studies that evaluated the completeness of information on GS and CS notification cases in Latin American and Caribbean countries^19^. This situation emphasised the need to improve the capacity of countries to collect high-quality data for diseases of interest to public health^7,19^.

GS and CS notifications have increased more than 9 and 5 times, respectively, in recent years, possibly due to the expansion of primary health care coverage, in particular the Family Health Strategy, and the Bolsa Familia Program, which conditions the receipt of the benefit to the attendance of pregnant women to prenatal care and children to medical appointments to monitor growth and development^14,20^. Improvement in case detection has not been followed by an increase in the completeness of information, with significant regional differences across Brazilian regions, which corroborate previous results described in the literature for other infectious diseases^12,14,21^. Problems with information completeness of SG and SC records may be related to differences in the level of commitment of health professionals with regards to the adequate registration of case notification records, the reduced number of health professionals in health services to complete a significant number of records, as well as the non-prioritisation of syphilis by health managers^7,12^. Considering that, often, notification is seen as a bureaucratic activity of secondary importance, it is essential to train health professionals on the need to generate quality information for epidemiological surveillance, ensuring the integrated flow of information between the responsible sectors, which can reduce differences at the area of residence and regional level in the quality of the information available in syphilis notification records^5,7,22^.

The care score completeness for CS was classified as regular for the indigenous group and good for the other racial categories, indicating possible difficulties in the monitoring of indigenous children and, consequently, in the adequate case notifications, in addition to the absence or insufficiency in the training of indigenous health teams^23^. Many factors may contribute to the incompleteness of information, such as the availability of time to complete the INR fields, considering the demands of the service, the existence of linguistic and cultural barriers, and difficulty in entering data onto SINAN by the base-centres and/or Special Indigenous Sanitary Districts, since besides not being notifying units, they do not interact with those in the municipal health system^13,23^.

It should be highlighted that the “partner’s treatment” variable had a very poor and poor degree of completeness for GS and CS, respectively. We supposed that this occurs because, since September of 2017, the Brazilian Ministry of Health modified the definition of CS cases by including the criterion “adequate treatment of the pregnant woman” and disregarding the information on the simultaneous “partner’s treatment”. Therefore, this variable has become a complementary and non-mandatory filling field in INR^24^. Insufficient completeness of the field “concomitant treatment of the partner”, influenced by the non-mandatory registration of the information, harms the monitoring of syphilis cases, mainly CS, which reduces the possibility to assess the real transmission scenario of syphilis and the quality of prenatal care^12,25,26^. Studies indicate that the incompleteness of SINAN information, particularly for CS, and the high underreporting of cases, limit the use of this system in studies of incidence and factors related to the growth of syphilis in Brazil^7,27,28^.

Moreover, many variables present in INR are considered important markers for the monitoring and epidemiological evolution of GS and CS in the country (i.e., partner’s treatment for CS) and support the adoption of therapeutic conduct of the disease (i.e., clinical classification and time of diagnosis of the disease). Other fields, such as the mother’s level of education and race/colour of the mother or child, are used as markers of social and racial inequalities related to infection and/or associated negative outcomes^4,7,28^, which highlighted the importance to improve quality data in SINAN-Syphilis. Also, we should highlight that the incompleteness of information related to education level had been observed in several HIS throughout the national territory, limiting its use in research^29^.

Data incompleteness directly reflects on the quality of information by not demonstrating the reality of the epidemiological profile of syphilis cases notified in the country^30^. This scenario complicates the planning of strategies and actions to prevent and control syphilis, contributes to the inefficiency of public policies to tackle the disease, and interferes in achieving the goals of eliminating and eradicating syphilis cases proposed by the WHO^9,14,30^. This situation becomes more complex when we consider regional, residential, and ethnic-racial variations in the information completeness scores in a country of continental dimensions, such as Brazil. Therefore, it is suggested that future studies analyse the factors associated with the incompleteness of data and its variations among different groups and regions, which will allow the adoption of strategies to reduce differentials in the quality of SINAN-Syphilis information.

Our results indicate that there has been an improvement in the degree of completeness of SINAN-Syphilis information during the study period, despite variations observed. We present a proposal to evaluate the quality of SINAN-Syphilis information, which is still little explored in the national scenario, building scores based on the weighted average of completeness indicators that are easily operationalised. There are some limitations related to the definition of weights for the weighted average method, based on the experience of specialists and not through the correlation structure present in the data obtained by the PCA. However, weighting is commonly used to compile health indicators and, in our case, produces results consistent with those obtained via PCA. This study only accessed the completeness of information, one of the parameters used to evaluate the quality of the information, not considering cases of underreporting or agreement of information, which are also important parameters to be considered when we evaluate the quality of HIS^3^.

## Conclusion

There have been improvements in the completeness of GS and CS notifications in recent years, mainly with the care score variables, compared to the socioeconomic variables, although with differences between Brazilian regions, ethnic-racial groups, and area of residence for both diseases. However, essential information to describe the epidemiological profile and monitor GS and CS still have poor completeness, compromising the planning of epidemiological surveillance and health care actions. We recommended promoting health teams’ training and adopting strategies by health managers, aiming at improvements in the complete filling of INR, given the prioritisation of other demands in health services. Lastly, health professionals and managers can incorporate indicators of easy compilation and interpretability for monitoring the completeness of SINAN-syphilis data, as proposed in this study, into the routine of evaluating the quality of information aiming at its improvement.

## Data Availability

All data produced in the present study are available upon reasonable request to the authors.

https://datasus.saude.gov.br/transferencia-de-arquivos/

## Contributions

GLO, JSG, AJFF, and MYI contributed to the concept and design of the study. GLO and JSG prepared the data and conducted the manuscript analyses. GLO, JGS, AJFF, and MYI prepared the preliminary versions of the manuscript and interpreted the study data. RML, AMC, CT, MASS, RLF, ESP, RA, IOS, LS, and MLB contributed to data interpretation and critically reviewed the manuscript. All the authors approved the final version and are responsible for all aspects of the study, including guaranteeing its accuracy and integrity.

## Supplementary material

**Table S1**. Analysis of the completeness percentage and scores according to the year, region, area of residence, and race/colour of the pregnant woman, considering gestational and congenital syphilis, SINAN-Syphilis, 2007-2018.

